# Inflammation and fibrinolysis in loiasis before and after ivermectin treatment: a biological pilot cross-sectional study

**DOI:** 10.1101/2024.08.29.24312769

**Authors:** Tristan M. Lepage, Narcisse Nzune-Toche, Lucie A. Nkwengoua, Hugues C. Nana-Djeunga, Sebastien D.S. Pion, Joseph Kamgno, Charlotte Boullé, Jérémy T. Campillo, Michel Boussinesq, Claude T. Tayou, Cédric B. Chesnais

**Author notes:** Correspondence, Institut de recherche pour le développement (IRD), 911 avenue Agropolis, 34000 Montpellier, France.

## Abstract

We assessed the impact of loiasis and its treatment with ivermectin on hemostasis and inflammation in 38 adults in Cameroon. Participants were divided into four balanced groups based on their *Loa loa* microfilarial densities. At baseline, a positive correlation was observed between microfilarial densities, neutrophils (p=0.012) and eosinophils (p<0.001). At day 4 following ivermectin administration, mean D-dimers significantly increased, from 725 ng/mL to 1,276 ng/mL (p=0.024). Mean eosinophils rose from 225/µL to 1,807/µL (p<0.001). C-reactive protein, fibrinogen, and alpha-1-globulin also increased significantly after treatment. Ivermectin treatment appeared to induce inflammation and pronounced fibrinolysis, indicative of coagulation activation.

## Background

Loiasis, caused by the filarial nematode *Loa loa* and transmitted from human to human by tabanids, is exclusively endemic in central Africa. In infected individuals, adult female worms release embryos called microfilariae (mfs) which circulate in the bloodstream according to a diurnal cycle, with densities occasionally surpassing 100,000 mfs per mL of blood (mf/mL).

Although loiasis is historically considered as a benign disease, recent findings suggest that microfilaremic individuals have a significantly shortened life expectancy [1]. In addition, studies in the Republic of Congo showed that high *L. loa* microfilarial density (MFD) was significantly associated with proteinuria [2], cognitive impairment [3], and increased vascular stiffness [4].

Eosinophilia exceeding 5,000 /µL is common among subjects with loiasis. Eosinophils, known to express tissue factor, may trigger blood coagulation [5], leading to thrombosis. Reports indicate that arterial and venous thrombotic events can occur in patients with parasite-related hypereosinophilia [6], with documented instances of thrombosis [7] and micro-emboli formation in capillary vessels [8], occurring both spontaneously and as part of post-ivermectin *Loa*-related serious adverse events (SAEs). Evidence from primate models suggests an association between post-ivermectin life-threatening encephalopathy and hemostatic dysregulation, characterized by degenerating mfs in small vessels associated with fibrin deposition, endothelial damages, and hemorrhages [9]. Similarly, humans experiencing post-ivermectin encephalopathy often exhibit conjunctival and retinal hemorrhages [10].

A potential dysregulation of hemostasis may contribute to the excess morbidity and mortality associated with loiasis. However, data on this phenomenon remain limited and unexplored in humans. The objective of the present study was to assess for the first time the impact of both *L. loa* MFD and its treatment with ivermectin (IVM) on human hemostasis and inflammation blood parameters.

## Methods

### Study design and participants

The study was conducted from July to December 2022 in two Health Districts of the Center Region of Cameroon (Mbalmayo and Awaé) where loiasis is highly endemic. Participants consisted of consenting adults aged 25 to 70 who had been screened for the presence of *L. loa* mfs. Exclusion criteria included pregnancy, concurrent pro-inflammatory conditions (such as infection, cancer or hematological disorder), current use of anticoagulant or anti-inflammatory medications, or known allergy to IVM. Eligible participants were stratified into 4 groups based on their *L. loa* MFD: 0 mf/mL; 20–4,999 mf/mL; 5,000–19,999 mf/mL; and 20,000–40,000 mf/mL.

Venous blood samples were collected from all groups at baseline (D0) to measure hematological, hemostatic, and biochemical parameters. Subsequently, individuals in the 20–4,999 mf/mL and 5,000–19,999 mf/mL groups received a single oral dose of IVM (150µg/kg), and the same blood parameters were reassessed four days after (D4) treatment.

### Sample size

No data exists on the relationships between loiasis and hemostasis. In a study evaluating D-dimer levels in dogs infected with another filaria (*Dirofilaria immitis*) [11], the proportion of elevated D-dimers (>0.2 μg/ml) was 48% (22/46) in microfilaremic dogs, compared to 0% in amicrofilaremic dogs. Based on this finding, the inclusion of 16 *L. loa* microfilaremic subjects and 16 amicrofilaremic subjects would provide an 80% power with an alpha risk of 5% to detect such a difference in loiasis. However, it is difficult to extrapolate these assumptions to loiasis, which is a different disease with a different host. Therefore, we planned to include 10 *L. loa* amicrofilaremic subjects, and 30 microfilaremic subjects in 3 groups of ascending MFD.

### Laboratory analyses

Blood was collected by fingerprick between 10:00 am and 3:00 pm in non-heparinized capillaries and spread on microscope slides to prepare two 50 µL thick blood smears which were dried, dehemoglobinized, and stained with Giemsa within 24 hours. *L. loa* mfs present on slides were counted using a microscope at 100-fold magnification at the Higher Institute for Scientific and Medical Research in Yaoundé, and averages of both slides were multiplied by 20 to express MFD per milliliter of blood.

Venous blood samples were collected from each participant using EDTA, sodium citrate, and dry blood tubes. Complete blood count, erythrocyte sedimentation rate (ESR) after one hour, C-reactive protein (CRP) levels, prothrombin time (PT), activated thromboplastin time, and fibrinogen levels were measured at the hematology laboratory of the Yaoundé University Teaching Hospital.

Wright-stained blood smears were prepared and the percentage of eosinophils was recorded after examining 200 leukocytes. Eosinophil counts were calculated by multiplying the percentage of eosinophils by the white blood cell count.

Serum protein electrophoresis, ferritin and D-dimers were assessed at the Centre Pasteur in Yaoundé. Finally, prothrombin fragment F1+2 and plasminogen activity were evaluated in a subset of participants (10 without microfilariae and 10 with 20–40,000 mf/mL) at the Cerba Laboratory in France.

### Statistical analysis

Hematological, hemostatic, and biochemical blood parameters were compared among the four MFD groups at baseline using the Kruskal-Wallis test. When the test yielded a p-value < 0.1, a Cuzick’s test for trend was performed. Since D-dimers levels can be influenced by age and sex, the relationship between D-dimer and MFD was also assessed using a linear regression, with adjustments on age and sex. Prothrombin fragment F1+2 and plasminogen, assessed in a subset of 20 participants, were compared between amicrofilaremic and microfilaremic subjects using the Wilcoxon rank-sum test. Finally, within the groups of individuals treated with IVM (20–4,999 mf/mL and 5,000–19,999 mf/mL), changes in hematological, hemostatic, and biochemical blood parameters were compared between baseline and D4 post-IVM using a Wilcoxon signed-rank test for paired observations.

### Ethical considerations

The study adhered to ethical guidelines outlined in the Declaration of Helsinki, version 13, and was approved by the Ethics Committee for Human Research of the Catholic University of Central Africa (N° 2022/022503/CEIRSH/ESS/MH) and by Yaoundé University Teaching Hospital (N° 4181/AR/CHUY/DG/DGA/CAPRC). Only individuals who provided informed consent were enrolled and assigned an individual code for data recording. Blood parameters results were shared with all participants.

## Results

Twenty males and 18 females aged between 28 and 67 years (median age: 43 years old) participated in the study.

Table 1 presents the blood counts and the inflammation and hemostasis parameters according to the participants’ *L. loa* MFD categories. There was a trend towards increased leucocytes, neutrophils and eosinophils counts with increasing MFD (p=0.011, p=0.013 and p<0.001, respectively, Cuzick’s tests for trend). The mean eosinophil count increased from 118/µl in patients with no mf to 1,079/µl in patients with a MFD beyond 20,000 mf/mL. Although the distribution of platelets varied significantly between groups, no trend was observed regarding MFD levels (p=0.055, Cuzick’s test for trend). Inflammation and hemostasis parameters did not significantly differ between groups. Linear regression analysis of D-dimers by MFD, adjusted on age and sex, did not show any significant associations. Mean ESR were consistently increased across all groups (normal values: < 20 mm/h). Gamma-globulin were also elevated in all groups, with means from 21 to 27 g/L (normal values: < 11 g/L). Clonal gamma-globulin expression was suspected on the serum protein electrophoresis in 3 patients, each in separate groups.

Eighteen individuals with an MFD between 20 and 19,999 mf/mL received a single dose of IVM and were re-examined at D4 (Table 2). Post-treatment, the mean MFD significantly decreased from 6,211 mf/mL to 2,972 mf/mL. Eosinophil count means rose from 225/µL to 1,807/µL (p<0.001). Various inflammation markers changed significantly: CRP, ESR, fibrinogen and alpha-1-globulin levels increased (p=0.038, p=0.010, p=0.002 and p=0.044, respectively), while albumin level decreased p=0.044). Among hemostatic parameters, mean D-dimers significantly increased, from 725 ng/mL to 1,276 ng/mL after IVM (p=0.024) with a median relative variation of +34.7% (interquartile range: 1.44-72.5%). Prothrombin fragment 1+2 mean increased from 315 to 435 pmol/L, but the difference was not statistically significant (p=0.313). No new clonal gamma-globulin expression on protein electrophoresis was suspected, while previously identified clonal expression in treated patients did not change after IVM.

## Discussion

In this pilot biological study, we describe for the first time hemostasis and inflammation parameters associated with loiasis. Our results show a remarkable trend towards increased leukocytes, mainly driven by elevated eosinophils and neutrophils, with increasing L. loa MFD. In a 2022 biological study in Gabon [12], eosinophil counts were also positively correlated with *L. loa* MFD, with an estimated increase in eosinophil counts every 10-fold increase in parasitemia (p-adj. = 0.012, ß-estimate: 0.17[0.04–0.31]). Chronic eosinophilia, a characteristic feature of loiasis, is known for its ability to trigger severe health conditions, such as endomyocardial fibrosis, tropical pulmonary eosinophilia, and thromboembolic events, consequently increasing the risk of mortality associated with loiasis [1].

The notable increase in neutrophils that correlates with increasing MFDs raises intriguing questions. Prior research has associated neutrophil recruitment with the presence of endosymbiotic *Wolbachia* bacteria in filarial infections (onchocerciasis and lymphatic filariasis) [13]. However, as *L. loa* mfs lack *Wolbachia* bacteria, alternative mechanisms must be considered to elucidate neutrophil involvement in loiasis.

Furthermore, our study highlighted a polyclonal elevation of gamma-globulins across all MFD groups, suggesting a systemic immune response. This phenomenon is consistent with a previous study in Ghana, were 75% of participants had gamma-globulins > 16 g/L [14]. This hypergammaglobulinemia might be explained by the influence of genetic as well as environmental factors. Mean ESR were increased in all groups, possibly driven by hypergammaglobulinemia, which is known to accelerate erythrocyte sedimentation.

Interestingly, while inflammation and hemostasis parameters did not significantly differ with *L. loa* MFD at baseline, treatment with IVM resulted in significant alterations in several inflammatory markers (such as CRP, ESR, fibrinogen and alpha-1-globulins). While post-IVM CRP increase has been reported previously [15], we document for the first time significant changes in a range of inflammatory parameters. We observed a marked increase in eosinophils, consistent with previous reports of interleukin-5 (IL-5)-driven eosinophilia following microfilarial killing by IVM [16].

The increase in D-dimers observed on D4 post-IVM suggests enhanced fibrinolysis and thus coagulation activation. This could be attributed to the close relationship between hemostasis and inflammation, where inflammation induces fibrin formation in the extravascular space, facilitating immune cells functions [17]. Consequently, secondary fibrinolysis generates D-dimers, which correlate with inflammation severity. However, this correlation is modest, as previous studies found a Spearman’s correlation coefficient of 0.18 between CRP and D-dimers, indicating a limited response of D-dimers to CRP elevation [18]. Therefore, in addition to inflammation, the potential involvement of thrombotic processes in the increase of D-dimers after IVM cannot be ruled out, potentially both driven by intravascular dying mfs and inflammation-induced hypercoagulable condition.

In addition to previous reports of post-IVM retinal emboli [10], this hypothesis is supported by autopsies of post-IVM encephalopathies in baboon models, which show abnormalities ranging from intravascular degenerating mfs and vessel wall infiltration by immune cells such as eosinophils, to fibrin deposition and endothelial disruption with extravascular hemosiderin deposits [9,19]. These findings provide a plausible pathological basis for the observed post-IVM treatment biological changes, potentially contributing to the pathogenesis of post-IVM SAEs.

Despite the valuable insights gained, our study has limitations. The small sample size and absence of preexisting data require cautious interpretation of our findings, highlighting the need for larger dedicated studies to validate our observations and understand their clinical implications. However, our results, particularly in baseline comparisons, may guide further research with adequate sample sizes, especially to explore the impact of loiasis on the hemostatic system. Additionally, potential confounding by other health conditions among participants, particularly in a low-resource healthcare setting, warrants consideration. Despite this, hematological changes showed a significant increase trend with *L. loa* MFD, aligning with previous findings [12]. However, as only *L. loa* MFD was assessed, potential confounding by other parasitic infections sensitive to IVM cannot be ruled out. Finally, our reliance on a single post-IVM evaluation timepoint may have limited our ability to detect biological abnormalities occurring outside this timeframe.

In conclusion, this pilot study identified several biological changes potentially associated with loiasis, occurring both spontaneously and after treatment with IVM. Further dedicated studies are needed to confirm these results, which might participate in loiasis excess mortality as well as post-IVM SAEs pathogenesis.

## Data Availability

Anonymized data will be hosted on the server https://dataverse.ird.fr/, and the terms of use will be those in force on the hosting site.

## Authors contributions

T.M.L, N.N.T., and L.A.N. carried out the cross-sectional study in 2021; C.B., M.B., C.T.T., and C.B.C designed the study; L.A.N. and C.T.T. performed hematological, hemostatic, and biochemical blood analyses; H.C.N.D. supervised parasitological analyses; N.N.T. and L.A.N. participated in the acquisition of data; T.M.L. and S.D.S.P. performed the statistical analyses; T.M.L. wrote the first version of the manuscript; H.C.N.D., C.B.C., M.B., S.D.S.P., C.T.T and J.T.C. reviewed the article. All authors approved the final version for publication.

## Acknowledgements

We thank all participants who agreed to take part in this study as well as the village chiefs and health workers involved.

